# Comparing human and artificial intelligence in writing for health journals: an exploratory study

**DOI:** 10.1101/2023.02.22.23286322

**Authors:** Zaeem ul Haq, Huzaifa Naeem, Ayesha Naeem, Faisal Iqbal, Durayya Zaeem

## Abstract

**Aim and objectives:** The aim was to contribute to the editorial principles on the possible use of Artificial Intelligence (AI)- based tools for scientific writing.

The objectives included

A. Enlist the inclusion and exclusion criteria to test ChatGPT use in scientific writing
B. Develop evaluation criteria to assess the quality of articles written by human authors and ChatGPT
C. Compare prospectively written manuscripts by human authors and ChatGPT

**Design:** Prospective exploratory study

**Intervention:** Human authors and ChatGPT were asked to write short journal articles on three topics: 1) Promotion of early childhood development in Pakistan 2) Interventions to improve gender-responsive health services in low-and-middle-income countries, and 3) The pitfalls in risk communication for COVID-19. We content analyzed the articles using an evaluation matrix.

**Outcome measures:** The completeness, credibility, and scientific content of an article. Completeness meant that structure (IMRaD) and organization was maintained. Credibility required that others work is duly cited, with an accurate bibliography. Scientific content required specificity, data accuracy, cohesion, inclusivity, confidentiality, limitations, readability, and time efficiency.

**Results:** The articles by human authors scored better than ChatGPT in completeness and credibility. Similarly, human-written articles scored better for most of the items in scientific content except for time efficiency where ChatGPT scored better. The methods section was absent in ChatGPT articles, and a majority of references in its bibliography were unverifiable.

**Conclusions:** ChatGPT generates content that is believable but may not be true. The creators of this powerful model must step up and provide solutions to manage its glitches and potential misuse. In parallel, the academic departments, editors, and publishers must expect a growing utilization of ChatGPT and similar tools. Disallowing ChatGPT as a co-author may not be enough on their part. They must adapt the editorial policies, use measures to detect AI-based writing, and stop its likely implications for human health and life.

**STRENGTHS AND LIMITATIONS:**

- First study that examines the scientific writing of ChatGPT by comparing it with human-written articles.
- Explains how ChatGPT generates believable content that may not be true.
- Indicates that the creators of ChatGPT must step up to address its misuse and potential hazards.
- An initial exploration, based on limited data—larger studies are needed for generalizable conclusions.

## INTRODUCTION

Evidence-based medicine (EBM) is a building block of modern clinical care and public health. EBM relies on sound evidence collected through rigorous experimental designs and their reported results [1,2]. The pooled results from these experiments are published in the form of systematic reviews, which public representatives and organizational heads use for policy formulation, and clinicians as well as public health physicians follow in their practice. Writing and publishing scientific data is an essential step throughout these processes.

Writing up scientific information demands professional integrity—writing about the science of human health requires added responsibility. The arrival of Generative Pre-trained Transformer (GPT) has opened new discussions about scientific integrity and responsibility. The latest version, i.e., ChatGPT is a language model that uses deep learning to write text like a human being writes it. Released on 30 November 2022 by OpenAI, ChatGPT is available to the public for free for a period of research preview [3]. When asked, ChatGPT can fetch internet data that statistically appear correlated, and put it together as a piece of writing.

Several authors have published their views after experimenting with ChatGPT. Dowling and Lucey have demonstrated that with due input from human authors and through three successive iterations, an argumentative article on economics could be refined, and all three iterations were publishable [4]. Gao et al. asked ChatGPT to write 50 abstracts already published in five top-rated health journals and found that ChatGPT could generate content without plagiarism [5]. A significant proportion (32%) of these abstracts evaded the detection by the human eye—reviewers marked them as written by human authors. Moreover, there have been instances where journals published papers with ChatGPT as a co-author [6,7].

Responding to the developments, medical editors and publishing platforms have declared that since ChatGPT cannot be held accountable for what it writes, it cannot be an author [8,9]. If authors use it, they will have to explicitly mention this use in their methods [10,11]. The decision implies that authors can include one or more portions written by ChatGPT in their articles, provided they mention its use in the methods section. How the editors and publishers will ensure the argument’s quality or the robustness of science in these circumstances is unclear.

Against this backdrop, two questions are important from the perspective of health sciences literature. One, how correct and credible is the information that ChatGPT presents. Two, how complete is a paper written by ChatGPT in terms of essential elements required for publishing in a health journal. To respond, we conducted this study to compare three prospectively written journal articles by human authors with three articles on the same topics generated by ChatGPT. In light of the findings, we discuss some fundamental questions.

## METHODS

Ours was an exploratory study in which we examined the suitability of ChatGPT for science writing. We compared newly written short articles by human authors and ChatGPT on three different topics. The aim of our study was to contribute to editorial principles on the possible use of Artificial Intelligence (AI) based tools like ChatGPT for scientific writing.

The objectives included

For inclusion and exclusion criteria, since ChatGPT can use only existing information and cannot create empirical data (although it can generate the report of a randomized controlled trial if asked), we decided to select topics that could be handled by reviewing the existing literature. Secondly, we decided not to compare ChatGPT outputs with published articles because an already published article would be available for replication. We, therefore, asked to write original articles. Third, we decided that shorter manuscripts could serve the purpose and set a limit of 500 words for each article. Fourth, we included the words “structured”, “journal article”, “citation”, and “bibliography” in our instruction to both human authors and ChatGPT. We did not expect tables or figures in such short articles; therefore, we did not include these in our command.

For evaluation criteria, we searched for checklists to assess the quality and comprehension of a journal article. The available guidelines on EQUATOR, e.g., CONSORT for randomized controlled trials and PRISMA for systematic reviews, were mainly focused on the quality of research and did not serve our purpose. The closest we could get for our study was the EASE guidelines, which address three major areas including *structure* (title, abstract, introduction, methods, results, discussion, and acknowledgments), *scientific content* (clarity of thought, cohesiveness, and specificity of responses), and *credibility* (distinguish own data from others, cite others’ work and add a list of references) [12]. After team discussions and initial testing, we finalized a 14-item evaluation matrix (Table 1) aligned with the EASE dimensions. All items had a Likert scale of 1-5, which two study team members – not involved in the writing part of the study – used to score the articles. The evaluators received blinded versions of the articles, and independently read and scored them. We used another AI-based tool, Grammarly, for scoring two items, i.e., originality and readability. Grammarly detects plagiarism and reports the unoriginal proportion of writing. It also generates a score about readability—the ease with which a student in eighth grade can read and understand a piece of writing.

**Table 1:** Evaluation matrix to compare human and AI-written structured articles on health.

| No. | Criteria | Definition | Scale | Objective of the item |
| --- | --- | --- | --- | --- |
| **1.** | **Completeness** |  |  |  |
| 1.1. | Structure | The writing follows the conventional (Introduction, Methods, Results, and Discussion, i.e., IMRaD) structure | 1-5 | To see whether the IMRaD sequence is there (with or without headings) |
| 1.2. | Organization | The writing logically flows from Introduction to Objective to Methods to Results and Discussion/conclusions. | 1-5 | To see whether the writing has a logical flow |
| **2.** | **Credibility** |  |  |  |
| 2.1. | Citations | The paper duly cites others' scholarly work, using a consistent citation style | 1-5 | To assess whether writing acknowledges others' work. |
| 2.2. | Bibliography | The paper includes a complete bibliography of all citations it made | 1-5 | To see the completeness of referencing |
| 2.3. | Credibility of references | The papers/studies cited exist and are not phantom | 1-5 | To confirm that references are authentic. |
| 2.4. | Originality | The proportion of plagiarism reported by Grammarly | 1-100 | To assess the originality of the work |
| **3.** | **Scientific content** |  |  |  |
| 3.1. | Specificity | The content provides specific answers to the assigned task/s | 1-5 | To assess how specific the outputs are to the assigned tasks |
| 3.2. | Data usage & Accuracy | Authors use numbers or qualitative data to make the argument, and the data is factual and accurate. | 1-5 | To rule out fictitiousness |
| 3.3. | Cohesion | The IMRaD sections have congruence to support the final argument | 1-5 | To evaluate the internal congruence of various sections |
| 3.4. | Inclusivity | If relevant, the article mentions what the argument means to different genders, and segments of the population | 1-5 | To assess the inclusivity of oft-ignored populations in papers |
| 3.5. | Confidentiality | The degree of confidentiality practiced to prevent the participants' identification | 1-5 | To assess ethical robustness |
| 3.6. | Limitations | The paper mentions study limitations | 1-5 | To assess the author's ability to acknowledge study limitations |
| 3.7. | Readability | Readability score generated by the writing software Grammarly | 1-100 | To assess the readability of the article by a wider audience |
| 3.8. | Time-efficiency | The amount of time (minutes) spent in completing the task | 1-5 | To assess time efficiency |

For objective C, we decided on three diverse topics, including “Promotion of early childhood development in Pakistan”, “Interventions to improve gender-responsive health services in low-and-middle-income countries”, and “The pitfalls in risk communication for COVID-19”. Our instruction read: “Write a structured article of 500 words with citations and bibliography on ….”. Two study team members wrote the human versions of these short articles parallel to ChatGPT. The outputs from human authors and ChatGPT were shared as MS Word Files 1 and 2, respectively, with evaluators who did not have a role in writing the short articles. They independently scored both categories of articles, and scores from both were combined to calculate averages. The Grammarly scores of originality and readability were divided by 20 to bring them at par with scores of other items in our list and make the entire data comparable. We used Excel to compile the results, calculate averages and draw the graphical representation.

The study was conducted in January-February 2023 and did not require ethical approval, as human subjects were not directly involved. We include the human and ChatGPT written articles in Annex 1 and Annex 2 respectively, in this paper.

## RESULTS

We present average scores by human authors and ChatGPT (Figure 1) on various items under a scholarly article’s three dimensions: *structure and organization, credibility*, and *scientific content*.

**Figure 1:**
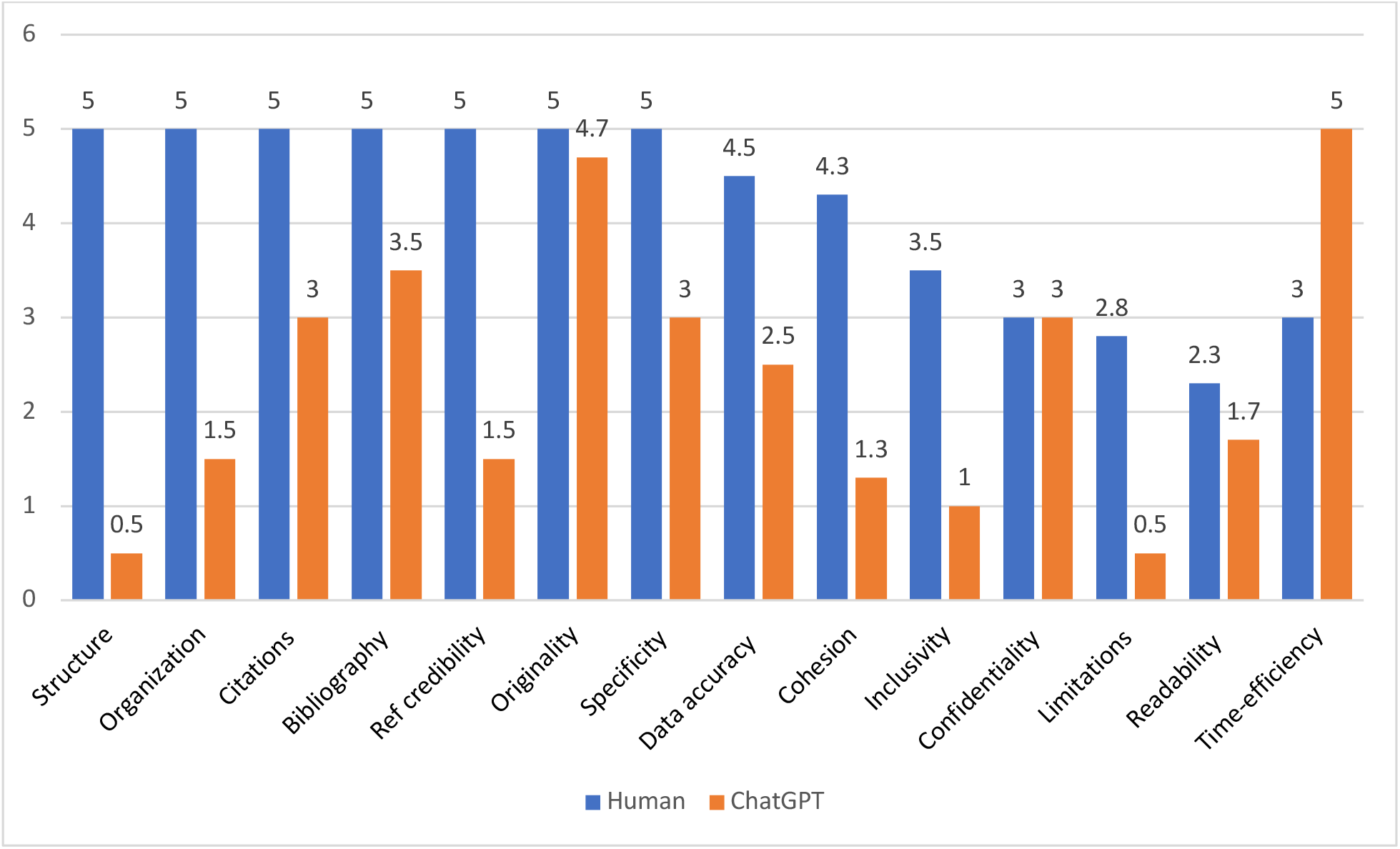
Comparative performance of human and ChatGPT outputs on different writing tasks.

### Structure and organization

Compared to the articles by human authors, which scored perfect, the ChatGPT articles did not score well. They did not have the IMRaD structure nor the logical flow of introducing the problem, highlighting the knowledge gap, describing the methods, presenting results, and ending with a discussion. Especially conspicuous was the absence of the “methods” section. After introducing the topic, all ChatGPT articles directly presented results mixed with discussion. All the articles started by introducing the topic, and all ended in a concluding paragraph that started with “in conclusion” making the writing look mechanical. Moreover, there was a distinctive void between results and discussion making it difficult to understand the premise of the discussion.

### Credibility

Compared to the articles written by human authors, which scored a perfect five, the citations and references part of ChatGPT articles was modest. The work from other scholars was cited in APA format, and a list of references was also provided at the end of the article that matched the citations. However, the citations were minimal in number and did not include even a single systematic review or journal article in the ChatGPT-generated articles. Only the grey literature, i.e., organizational reports or documents, was included. Moreover, the URL for these web-based documents was absent. When we searched for these references, we could not find most of these documents. Additionally, some references appeared a fabrication of different words put together as a report title.

### Scientific content

Grammarly detected no or negligible plagiarism in both types; therefore, both had a perfect or near-perfect score in originality. The specificity of the response to the given task was perfect for the human written but moderate by the ChatGPT written pieces. The absence of “methods” created a vacuum in which the presented information seemed weak leading to a moderate specificity score. Moreover, the evaluation team thought the writing was mechanical and devoid of an insightful discussion. The data usage and accuracy for ChatGPT articles scored low compared to the human-written articles. This low score by ChatGPT was due to the absence of numerical data in all three articles. The overall writing appeared more cohesive in the human-written articles than in those generated by ChatPT. The inclusivity score was better for human authors compared to ChatGPT, as was the score on study limitations. The confidentiality was not relevant as these articles were not based on empirical data. Both human-written and ChatGPT articles scored low on readability item. For efficiency, the ChatGPT articles had a perfect score. The time consumed in writing three pieces was about five minutes, whereas the human authors took one week to write those short articles.

## DISCUSSION

At the time of writing, this is the first study that examines the potential of ChatGPT in prospective writing for health journals by comparing it with human-written articles. With Chat-GPT, using AI in different scholarly tasks, including writing, is being widely discussed. We find that while ChatGPT can efficiently produce articles that appear original and coherent in their argument, a deeper examination reveals they are perfunctory. The “methods” part, which provides a scientific basis for an argument and opens chances of replicability by other scientists, is entirely missing. Moreover, the scholarly work ChatGPT quotes may not exist. This weakness puts a question mark on the accuracy and reliability of the entire writing.

Authors in health have raised similar issues while examining different types of scientific content. Spitale et al. who studied an earlier version, i.e., GPT-3, found that the GPT-3 generated tweets on health issues have more accurate and understandable information than tweets by a human being [13]. GPT-3 could also generate disinformation that was more compelling. Importantly, in their study, humans could not distinguish tweets generated by GPT-3 from tweets from humans. In another study, Gao et al. published similar findings that ChatGPT could write original abstracts, with 1/3^rd^ evading the human detection of the authorship [5]. They concluded that ChatGPT could write believable scientific content, though with wholly generated data. The content is devoid of plagiarism, but AI output detectors and careful human reviewers could mostly detect it.

However, none of these studies examined the citations in the generative content: one examined abstracts, which do not contain citations as a rule and the other studied tweets. Bloggers and commentators, however, have alluded to the potentially misleading or inaccurate content that ChatGPT can produce and may cause harm [14,15]. The apprehension becomes manifold in the background of the infodemic — much of which was misinformation—during the pandemic of COVID-19. Instances have been reported where an author found the references to be incorrect, and when questioned back, ChatGPT “excused” that the reference did not exist and that it had made a mistake [14].

Authors like Dowling and Lucey, who have a focus on finance research and not health, have found ChatGPT outputs promising. They adopted a three-step iterative approach to develop and refine a study using ChatGPT. They argue that ChatGPT can help improve a research idea, create a dataset, conduct a literature review, and give suggestions for testing and examination [4]. Others have praised ChatGPT for being socially responsible on questions about which they as authors expected a biased response but were surprised by a balanced statement [16]. A recent *Lancet* editorial also suggests that ChatGPT use could be rationalized not for scientific content but for higher readability [11]. In our case, however, in addition to the unverifiable references as a weakness, we also found that ChatGPT content did not earn a readability score higher than human authors.

Our study has limitations as it is an exploratory study that provides some evidence about the unscientific approach and unauthentic data put forth by ChatGPT. More research is required to have a complete picture. In addition to the limited data we could utilize, this study was conducted in an evolving environment in which ChatGPT is also undergoing changes and may bring relevant improvement in its functionality. However, it is also a strength of our study because the improvements in such technologies become possible with their utilization and sharing of experiences.

The scholarly discussion so far has been on publication ethics and the possible monetization of ChatGPT that may exacerbate the existing knowledge inequity [15,17]. Our concern is the lack of accuracy in information – ChatGPT’s capability of putting together any information in a manner that makes it believable – and the likely harm it can cause to human life and health. If ChatGPT can create flawed references, what about the information for which it is using these references? The potential harm such technologies can cause because of producing misleading or inaccurate content must be considered before moving to other issues and discussions [18].

Spitale et al. have proposed that until we do not have efficient strategies for identifying disinformation (whether based on human skills or future AI improvements), it is better to restrict the use of technologies like ChatGPT. According to them, waiting is better till OpenAI has critically evaluated the implications and taken action to mitigate any adverse effects on society [13]. Along the same lines, and pinning the responsibility on OpenAI, others have said that the creators of such a powerful model, are also responsible for stepping up and providing solutions to manage its potential misuse [16].

However, it is also true that the genie is out of the bottle, and academic departments, editors, and publishers must expect a growing utilization of ChatGPT and more such tools. They must respond to the situation and make appropriate adjustments to the existing editorial policies [19]. Considering our study, we feel the science community should have a wide-ranging debate where they share experiences of using AI and reach a consensus on future actions. Authors must show responsibility, honesty, integrity, and transparency [17]. In parallel, the academic departments, editors, and publishers must expect a growing utilization of ChatGPT and similar tools. Disallowing ChatGPT as a co-author may not be enough on their part. They must adapt the editorial policies, use measures to detect AI-based writing, and stop its likely threats for human health and life.

## Supporting information

Annex 2

Annex 1

## Data Availability

All data produced in the present work are contained in the manuscript.

## Contributors

ZH developed the concept and design of the study and generated ChatGPT outputs. DZ and HN wrote the human part of the articles. AN and FI evaluated all the articles. ZH and DZ wrote the first draft. All the authors critically revised the manuscript and gave the final approval of the version to be published.

## Funding

This research received no specific grant from any funding agency in the public, commercial or not-for-profit sectors.

## Competing interests

None declared.

## Patient and public involvement

Patients and/or the public were not involved in the design, or conduct, or reporting, or dissemination plans of this research.

## Notes

### Competing Interest Statement

The authors have declared no competing interest.

### Funding Statement

This study did not receive any funding.

