## Supplementary material for "Comparing human and artificial intelligence in writing for health journals: an exploratory study": Annex 1

**Promoting Early Childhood Development: The Policy and Program Situation in Pakistan**

### Introduction

Early Childhood Development (ECD) is the process of maturation of a child's sensory, motor, cognitive, language, socio-emotional, and self-regulation skills (Richter et al., 2016). An optimal development during the early years of life warrants satisfactory schooling and enhanced productivity during adulthood—everything paving the way for sustainable development (Daelmans et al., 2016; Richter et al., 2016). Pakistan is a lower-middle-income country (LMIC) that has a high fertility rate. Moreover, about 64 million (29%) of its existing population is below 10 years of age (Government of Pakistan, 2017). A large proportion of Pakistan's population lives below the poverty line (World Bank, 2022). Focusing on its children and youth to build the foundations of a productive society must be a high policy priority. In this paper, we examine the policy and program situation for ECD in Pakistan.

### Methods

We carried out a multistage literature review. Starting with an appraisal of global literature, we defined ECD and identified Nurturing Care Framework (NCF) to mark five essential dimensions of an ECD promotion program. These included health, nutrition, responsive caregiving, security and safety, and early learning opportunities (World Health Organization, 2018). The NCF also guides programs to cover three age groups: conception–two years, 3–5 years, and 6–8 years. We discussed these NCF-recommended dimensions and the age groups with experts and officials from the health, education, and social protection sector in Pakistan to reach a consensus about the definitions. In the second stage, we obtained published and grey literature on ECD in Pakistan and examined it for the availability of policy guidance on five areas of NCF for all three age groups of children. Two members of the research team independently examined the documents and discussed their findings to reach a consensus.

### Results

The ECD landscape in Pakistan is fragmented, with no shared understanding of the concept (Alam et al., 2022). The devolution of key functions following the 18th amendment to the Constitution, further adds to the challenge. While strong policy and service delivery networks exist for nutrition, the same is not true for health and education (Zaidi et al., 2018). Both these i.e., health and nutrition are not fully integrated, nor do they necessarily seek to address child development from a whole-of-early-childhood perspective (Zaidi et al., 2018). Other relevant sectors, such as child protection, social protection, disaster risk management, and water and sanitation, are not coordinated through an ECD framework. Moreover, key components of the NCF, like maternal mental healthcare, support to families to provide responsive caregiving, and support for early learning in the home are not addressed. Existing and new service delivery platforms may be leveraged for effective ECD provision. Presently, however, this is not the case in the absence of a coherent policy (Zaidi et al., 2018).

### Discussion and Recommendations

Pakistan needs an ECD policy that draws on global learning and guidance like the Nurturing Care Framework. Clear and unified definitions with costed strategies are required that take a holistic and equity-focused approach. The policies must be sufficiently flexible to facilitate their application in all sub-national units. Responsive parenting is a largely neglected area even among educated, urban families, and requires immediate attention. Addressing maternal mental health, screening for infant disability, and accidents affecting children is also important. Addressing the development needs through all stages of early childhood, including 5-8 years, as well as support for caregivers and families is of utmost importance. Finally, the policy must have an equity lens to reach all children regardless of gender, socioeconomic status, geographical location, or nomadic status, and factors like religion, caste, ethnicity, linguistic or tribal affiliation, cultural background, and vulnerability to insecurity and natural disasters.

**Interventions to improve gender-responsive health services in low- and middle-income countries**

### Introduction

Despite the overall gains in Maternal, Newborn, and Child Health (MNCH), some countries still have high maternal and child mortality (Foreman et al., 2018; Yaya et al., 2021). Gender disparity is a common determinant in such societies (Yaya et al., 2021). Moreover, the health systems continue to ignore the Sexual and Reproductive Health Rights (SRHR) and other health issues of adolescent girls, adding to the already enormous burden of disease (Melesse et al., 2020). The broader health sector recognizes gender inequality and intersectional power dynamics as a driver of poor health outcomes, particularly for women and girls, and recognizes a corresponding need to respond with a transformative approach that addresses the root causes of gender inequality (Heise et al., 2019). However, not much evidence is available on how to make these transformative changes.

### Methods

We carried out a desk review of global literature to draw specific guidance. Our objective was to address the knowledge gap by providing practice-based evidence on making health systems gender-responsive in low-and middle-income countries (LMICs). We searched three health and social science databases including PubMed, Google Scholar, and Scopus during the second week of January 2023 for articles from the years 2005-2022. We used a variety of keywords and their combinations to search literature on how to improve the gender responsiveness of health systems in LMICs. In addition to research publications, we also explored grey literature in the form of policy documents and program guidelines for LMICs. From an initial pool of 18,689, we selected 48 articles that fulfilled our inclusion criteria. Using a specially developed review template for this purpose, two members of the team read the articles to find interventions that helped the health systems become gender responsive in LMICs. The reviewers then met and discussed their findings to reach a consensus.

### Results

Three sets of interventions emerged through this process. First, the Social and Behavior Change (SBC) interventions to improve household MNCH behaviors and healthcare utilization of women and young girls along with men's involvement and community support (George et al., 2019). Two, the responsiveness of the health system with improved infrastructure, equipment, logistics, and above all, the attitudes of health providers and their leaders towards women and young girls is crucial (Hay et al., 2019). Last, creating data systems by collecting information about women and young girls from the community to help the visibility of true magnitude of the problem, and planning for it accordingly (Plan International Canada, 2021).

### Discussion

Programs should direct their activities to SBC, service delivery, and data systems. Improving the visibility of the problem through data (unreported women and adolescent girls in this case), advocating with the system to acknowledge this problem and do something about it, and sensitizing individuals and the community to play their role in addressing this issue, is an effective strategy. Involving the community, especially women, in the management of their health facility, improves the quality and coverage of health services. We recommend that programs, implemented in a real-world setting, publish their findings to broaden the field of practice-based evidence for greater improvement in gender-responsive programs. This review is a preliminary exploration because it had time and resource constraints. Additional and expanded reviews are needed to broaden this knowledge.

**The pitfalls in risk communication for COVID-19**

**Introduction**

The world has entered the third year of COVID-19. Still, there are no signs of the pandemic ending any time soon (World Health Organization, 2022). Several evaluations of the response to this pandemic have been carried out both at international and country levels. Failure of risk communication is a consistent theme in these evaluations. Points supporting this view include: 1) Global agencies and the country governments could not build and maintain trust in their steps, 2) There was a lack of consistent and coherent messaging, and 3) Governments failed in addressing misinformation (Organization for Economic Cooperation & Development, 2022; Sachs et al., 2022). COVID-19 is not the first pandemic, nor will it be the last. Examining the pitfalls in RC strategies – an important component of response – is critical for improving the response to the present as well as future outbreaks. In this short paper, we review the global RC efforts and suggest modifications.

**Methods**

We carried out a quick literature review focusing on the evaluations of the COVID-19 response, and the published literature on RC. We searched health and social science search engines using the keywords, "risk communication", "community engagement", "crisis communication", "infodemic management", and "debunking", all combined with "COVID-19". Being a member of several international panels on RC, one of the authors had information from and access to the risk communication discussions and developments at the global level. This helped in identifying unpublished materials. We decided to include only those articles that focused on the performance of RC and included it in the title. Two reviewers independently read the articles and picked up themes that converged on to the focus of our study. Following this, they met and finalized the findings.

**Results**

With overall preparedness at 63% and RC capacity at 60% of the required level, the world struggled to respond when SARS-CoV-2 emerged from Wuhan, China in late 2019 (World Health Organization, 2020). The evolving situation demanded that experts admit a lack of full knowledge at the time they were issuing advice. Scientists and policymakers in many countries found it difficult to admit their incomplete knowledge and feared a loss of authority while issuing preventive advice (Smith et al., 2020). Moreover, they were confronting a highly politicized environment in which, unsubstantiated, but emotionally appealing misinformation was rampant (Wang et al., 2022), while quick remedies to neutralize this disinformation were not available. There was a tendency to rely on high officials in crisis situations, who usually came to respond when the problem had already escalated (Organization for Economic Cooperation & Development, 2022). Finally, not enough strategic guidance was available to tailor communications to the successive waves in the same pandemic (Haq et al., 2021).

**Discussion**

Our study reveals that often, the confusion was not in the communication, but in the policy process. The solution is that communication should be on the decision table. Secondly, it is the handling of issues –the pandemic included – by the authorities that lead to a lack of trust and partisan criticism, which RC cannot deflect. The most recent example is China, where 2.02 million confirmed cases of COVID-19 were reported till 13 January 2023. However, on 22 January 2023, the chief epidemiologist of China CDC said that 80% of the Chinese population was already infected and there was no chance of massive COVID-19 wave around the Chinese New Year (Reuters, 2023). This type of conundrum can only be prevented by being truthful all along and. Lastly, there is an urgent need of research to find effective techniques for making people capable of identifying misinformation and not mindlessly forwarding it(van Bavel et al., 2020). Testing of more theories for promoting prosocial behaviors is also required.
